# Bibliometric analysis of disaster nursing based on CiteSpace

**DOI:** 10.1101/2024.09.21.24314133

**Authors:** Haixia Xie, Zhi Zhang, Ruijuan Han, Fulan Li, Tianshuang Yu

## Abstract

**Background:** Disaster nursing as a response to natural disasters, accidents disasters and other emergencies important areas, on a global scale in recent years received wide attention and research. This study discusses the research status and development trend of disaster nursing in the past 10 years, to provide a reference for future research and practice.

**Methods:** Literature in disaster nursing from 2013 to 2023 was searched using the Web of Science database, and CiteSpace 6.1.R6 software was used for visual analysis.

**Results:** A total of 4865 articles were retrieved, identified, and presented with a trend of steady rise. The United States has the largest number and has significant influence in the field. Disaster preparedness, disaster resilience, and disaster nursing education are the research hotspots. The combination of disaster nursing and intelligent technology will be the future research trend.

**Conclusion:** In the last 10 years, research on disaster nursing has shown a diversified and comprehensive development trend. In the future, with the continuous impact of global climate change and the increasing social demand for disaster nursing, the research and practice of disaster nursing will pay more attention to interdisciplinary cooperation, technological innovation, and humanistic care.

## 1. Introduction

With the aggravation of global climate change and the frequent occurrence of natural disasters, the importance and urgency of disaster nursing as a comprehensive discipline have become increasingly prominent. In recent years, more and more scholars and experts have invested in disaster nursing research, which has made important contributions to responding to disaster challenges, improving nursing quality, and ensuring people’s safety. In the field of disaster nursing, the research hotspots and focus issues not only reflect the academic frontier and development direction of this field but also reflect the urgent needs and challenges in practice. Through bibliometric analysis, we can systematically sort out and analyze the research results in the field of disaster nursing and reveal its research characteristics and rules so as to better grasp the development trend and future direction of disaster nursing. Therefore, this paper aims to conduct a quantitative analysis of disaster nursing research literature from 2013 to 2023 through the Web of Science database and reveal the research hotspots and development trends of disaster nursing in the past 10 years in order to provide valuable reference and enlightenment for future research and practice.

## 2. Methods

### 2.1 Data collection

We used the Web of Science Core Collection (WOSCC) advanced search feature, the only database that fully utilizes CiteSpace’s functionalities, to find relevant articles on “disaster nursing” (Chen, 2006). The search formula was TS=(“disaster nursing” or “disaster care” or “disaster nursing care”).The search selected English literature and articles and excluded proceedings papers, book chapters, data papers, and retracted publications. The study period was January 1, 2013, through December 31, 2023. CiteSpace version 6.1.R6 was employed. These data were independently verified by two authors. We all can not access to information that could identify individual participants during or after data collection

### 2.2 Data export

After eliminating any literature that is not relevant to the topic, we extracted the remaining 4865 articles using the Web of Science (WOS) platform in the format of “full records and cited references.” The data from the first to the 500th article were saved in a file named “download_1-500,” while the data from the 501st to the 1000th article were saved in a folder named “download_501-1000.” All data were exported in accordance with this pattern until 4865 articles were obtained.

### 2.3 Bibliometrics and visualization analysis

After removing duplicates and importing all WOSCC data into CiteSpace (version 6.1.R6), we converted the data into a format for visual analysis. CiteSpace utilizes nodes to indicate information such as country, institution, author, and keywords. In the graph for visual analysis from January 1st, 2013 to December 31st, 2023, “ Years Per Slice “ is set to 1 year. The size and color of the nodes, respectively, indicate the frequency and year of occurrence. The node radius increases as the node’s total frequency increases. The line between nodes represents the strength of the cooperation or occurrence between two nodes. Nodes with a red border are typically thought to have high centrality (centrality≥0.1), making them hot research topics and key positions in the field [1].

## 3. Results

### 3.1 Analysis of the annual output of publications

The quantity of academic papers published each year can provide insight into this field ‘ s past and forecast its future growth and decline. After using CiteSpace to eliminate duplicates, the number of published years was used as the horizontal axis and the number of published articles as the vertical axis to create a chart for the annual outputs of publications, as shown in Fig 1. This figure demonstrates that the amount of disaster nursing literature has fluctuated over time. In general, research interest in disaster nursing has increased significantly over the past decade, with the number of publications increasing from 243 in 2013 to 591 in 2023. The year with the fewest articles published was 2013 (243, 4.9%), whereas the number of publications peaked in 2021 (765, 15.7%). The reason may be that the novel coronavirus pneumonia swept the world at the end of 2019, bringing huge losses to human society and making the public once again aware of the importance of disaster nursing, so disaster nursing-related research has attracted global attention, and with the unremitting efforts of experts and scholars, international disaster nursing research has been continuously expanded.

**Fig 1.**
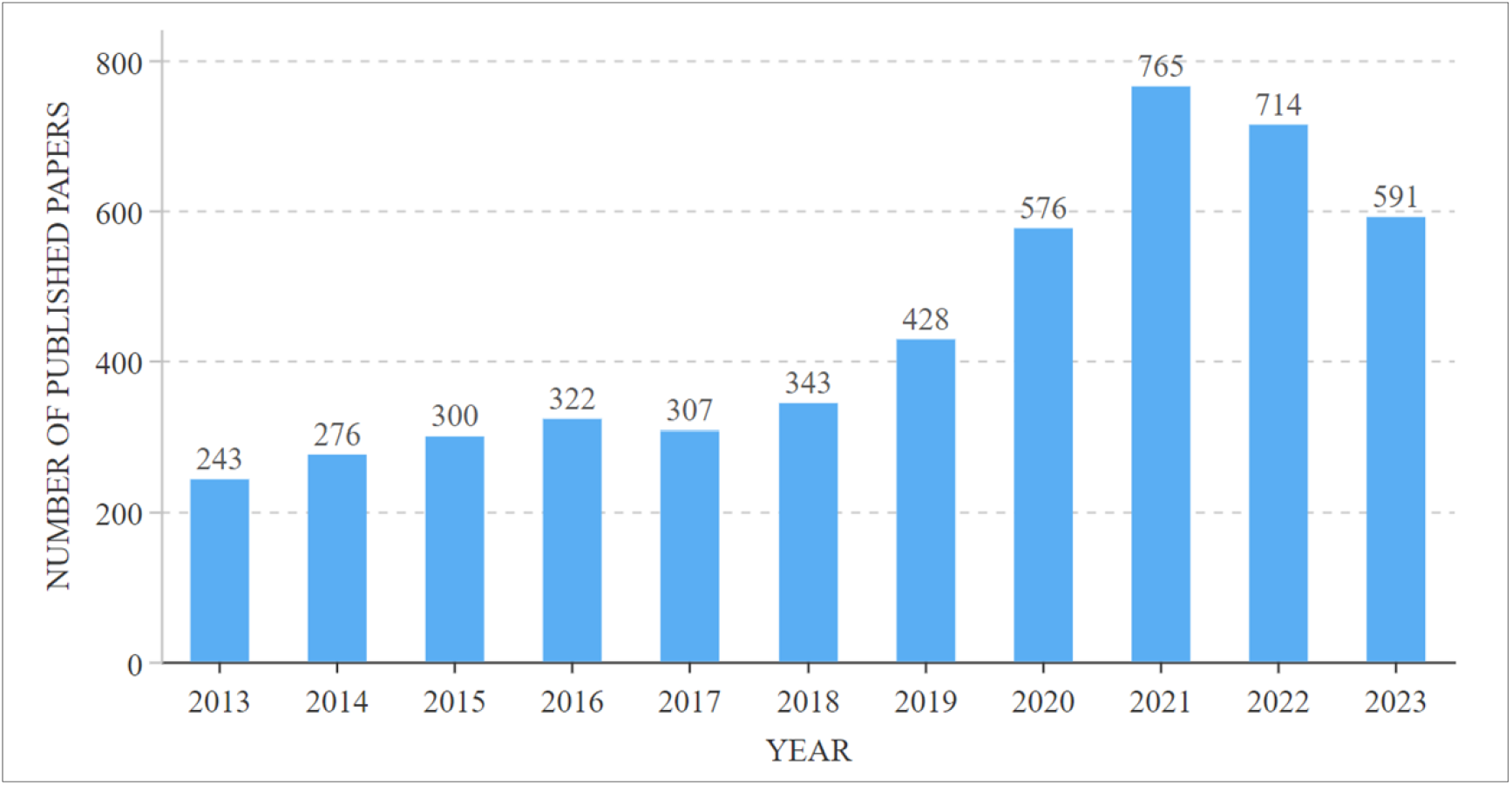
Annual chart of publications.

### 3.2 Analysis of countries or regions

The citation analyzer of the WOSCC database is used to count the number of documents sent by countries or regions, and the default setting of CiteSpace software analyzes the cooperative relationship between countries and regions. Fig 2 illustrates that over the past decade, 152 countries worldwide have conducted relevant research, with the United States leading the way with a total of 2202 papers, demonstrating its exceptional strength in scientific research. Table 1 displays the top 10 countries. The top five nations were the United States(2202,45%), Australia (405,8.3%), Japan (399,8.2%), England (369,7.5%), and China (345,7.0%). It can be seen that investment in disaster care research is mainly in coastal, economically developed, and disaster-prone countries, while the scientific research capacity of countries with lagging development, frequent disasters, and remote areas is obviously weak.

**Table 1.** Top 10 countries with publications of disaster nursing from 2013 to 2023.

| Rank | Countries | Counts | Centrality |
| --- | --- | --- | --- |
| 1 | USA | 2202 | 0.05 |
| 2 | Australia | 405 | 0.00 |
| 3 | Japan | 399 | 0.00 |
| 4 | England | 369 | 0.05 |
| 5 | China | 345 | 0.05 |
| 6 | Canada | 272 | 0.10 |
| 7 | Italy | 162 | 0.03 |
| 8 | Sweden | 152 | 0.00 |
| 9 | Iran | 143 | 0.00 |
| 10 | Germany | 126 | 0.22 |

**Fig 2.**
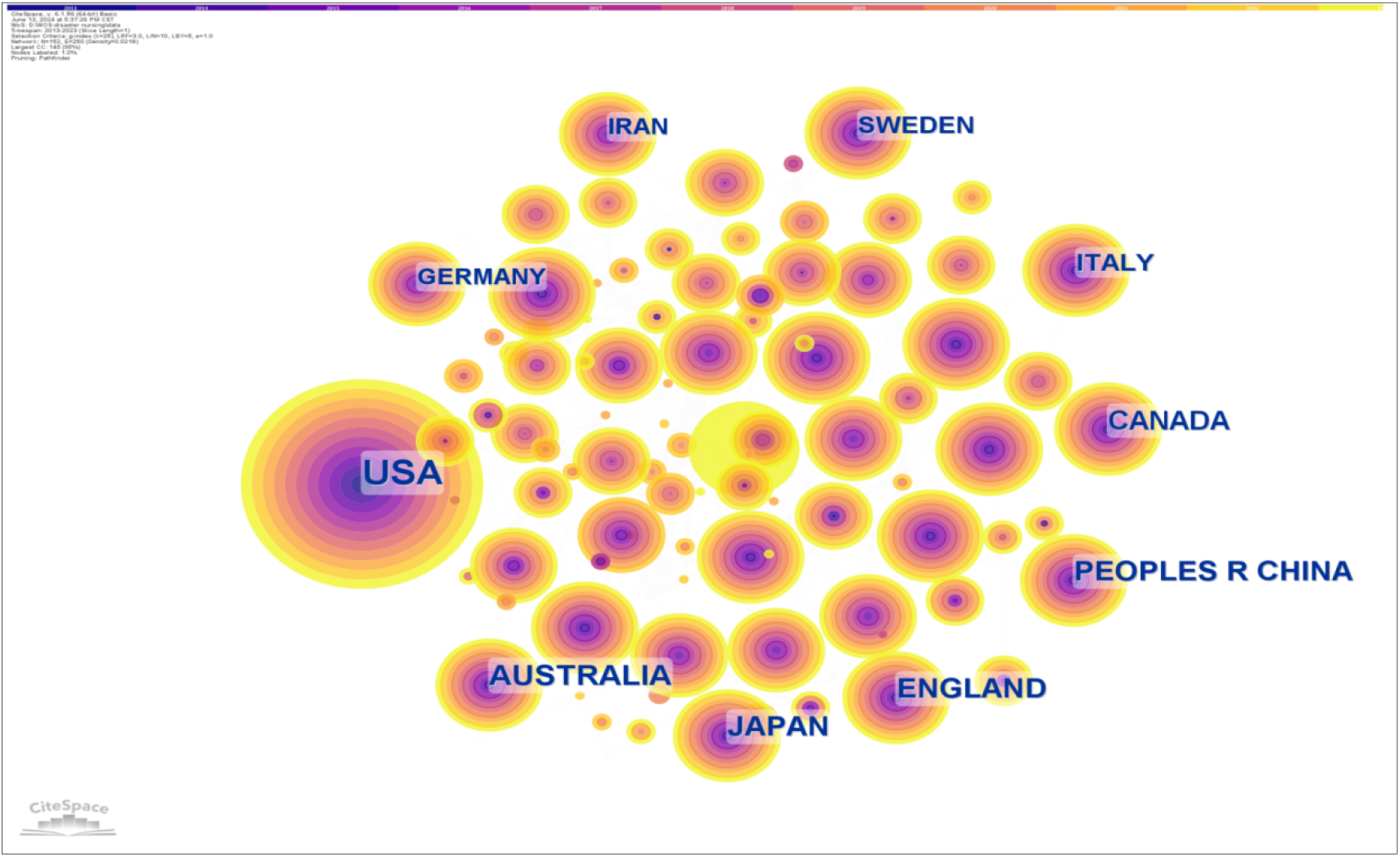
Map of countries.

### 3.3 Institutions

Table 2 provides information on the top 10 institutions that published the analyzed articles. The displayed data is an outcome of the default settings of CiteSpace software. These included 6 American institutions, 2 Japanese institutions, 1 Swedish institution, and 1 Australian institution. The node size indicates the number of documents sent, and the lines state the interagency cooperation, as shown in Fig 3.

**Table 2.** Top 10 institutions with publications of disaster nursing from 2013 to 2023.

| Rank | Institution | Count | Countries |
| --- | --- | --- | --- |
| 1 | Columbia University | 97 | United States |
| 2 | Harvard Medical School | 82 | United States |
| 3 | Johns Hopkins University | 81 | United States |
| 4 | Fukushima Medical University | 79 | Japan |
| 5 | Harvard University | 75 | United States |
| 6 | Tohoku University | 68 | Japan |
| 7 | New York University | 66 | United States |
| 8 | Karolinska Institute | 59 | Sweden |
| 9 | Uniformed Service University of the Health Sciences | 58 | United States |
| 10 | Monash University | 57 | Australia |

**Fig 3.**
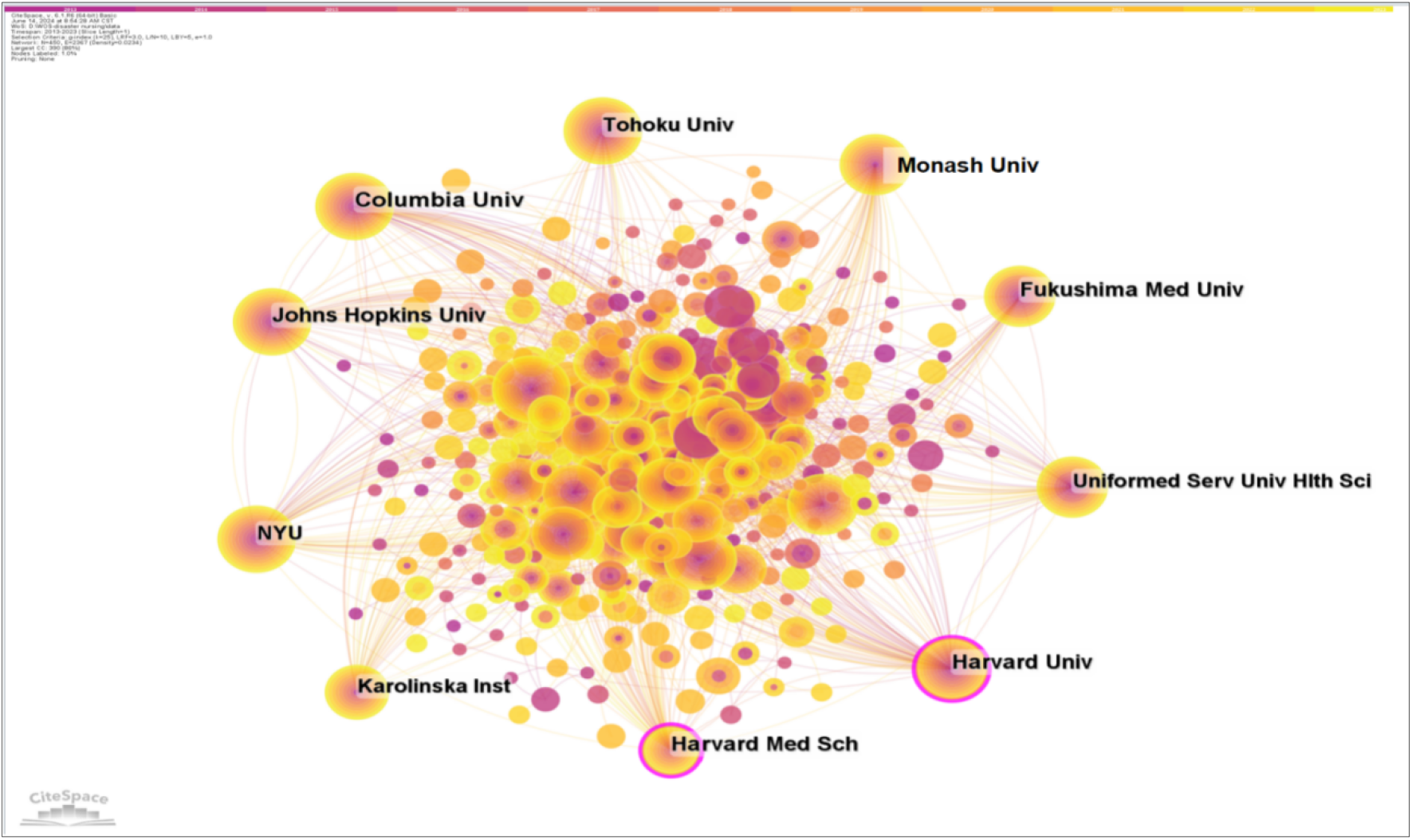
Map of institutions.

### 3.4 Analysis of authors

The nodes represent authors and lines represent cowork relationships in Fig 4. There are 549 nodes and 1,004 connections in the map, with a stable cooperative relationship formed between researchers. The top 10 authors in the publications, as indicated in Table 3. The most prolific author was Dobalian, Aram, who contributed 36 articles, followed by Tsubokura,Masaharu (29). Burkle,Frederick M had published 26 papers, had the highest centrality (0.10), and had strong cooperative relationships with other researchers. Nevertheless, the centrality of these high-yield authors’ nodes was low, indicating that they and other researchers had to establish more cooperation and communication to produce better research findings.

**Table 3.** Top 10 authors with publications of disaster nursing from 2013 to 2023.

| Rank | Authors | Count | Centrality |
| --- | --- | --- | --- |
| 1 | Dobalian,Aram | 36 | 0.00 |
| 2 | Tsubokura,Masaharu | 29 | 0.01 |
| 3 | Burkle,Frederick M | 26 | 0.10 |
| 4 | Della corte,Francesco | 23 | 0.01 |
| 5 | Ragazzoni,Luca | 23 | 0.07 |
| 6 | Ozaki,Akihiko | 18 | 0.00 |
| 7 | Khorram-manesh,Amir | 17 | 0.01 |
| 8 | Der-martirosian,Claudia | 17 | 0.00 |
| 9 | Ranse,Jamie | 17 | 0.00 |
| 10 | Christian,Nichael D | 15 | 0.01 |

**Fig 4.**
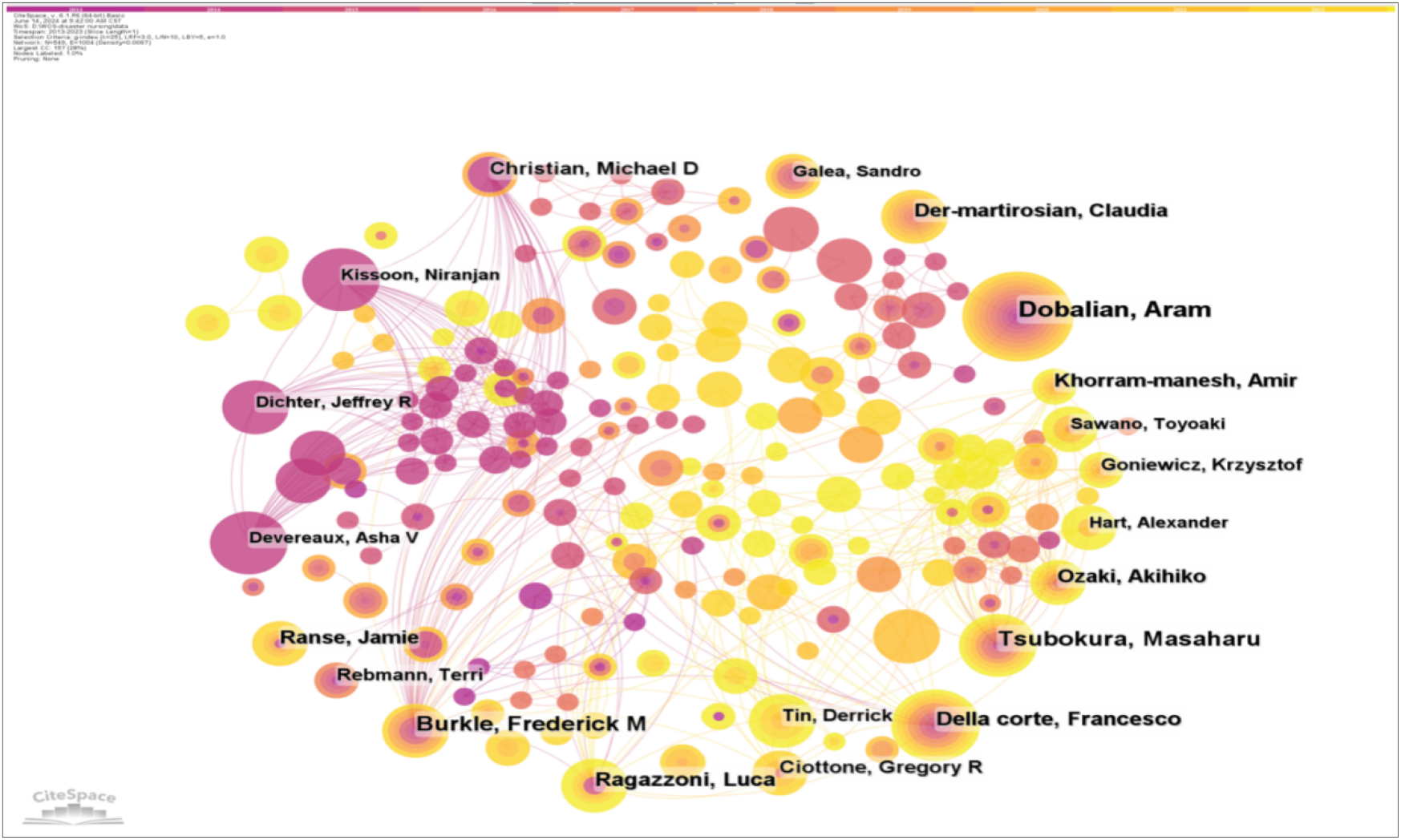
Map of coauthors.

### 3.5 Analysis of journals: overlay map

Fig 5 was created using CiteSpace’s “overlay maps” tool. The cited journals on the left and the cited journals on the right illustrate the links between them. The curve shows the citation relationship. The elliptic curve reflects the proportion between authors and publications. The more writers there are, the longer the ellipse’s horizontal axis is; as the vertical axis of the ellipse lengthens, more papers are published in the journal. Medicine, medical, clinical, neurology, economics, politics, psychology, education, and health were the key research sources for this subject. Publications related to health, nursing, medicine, molecular biology, genetics, education, social, economic, and political topics were widely cited. This suggests that the problem of disaster nursing is tied to medicine, education, economics and politics.

**Fig 5.**
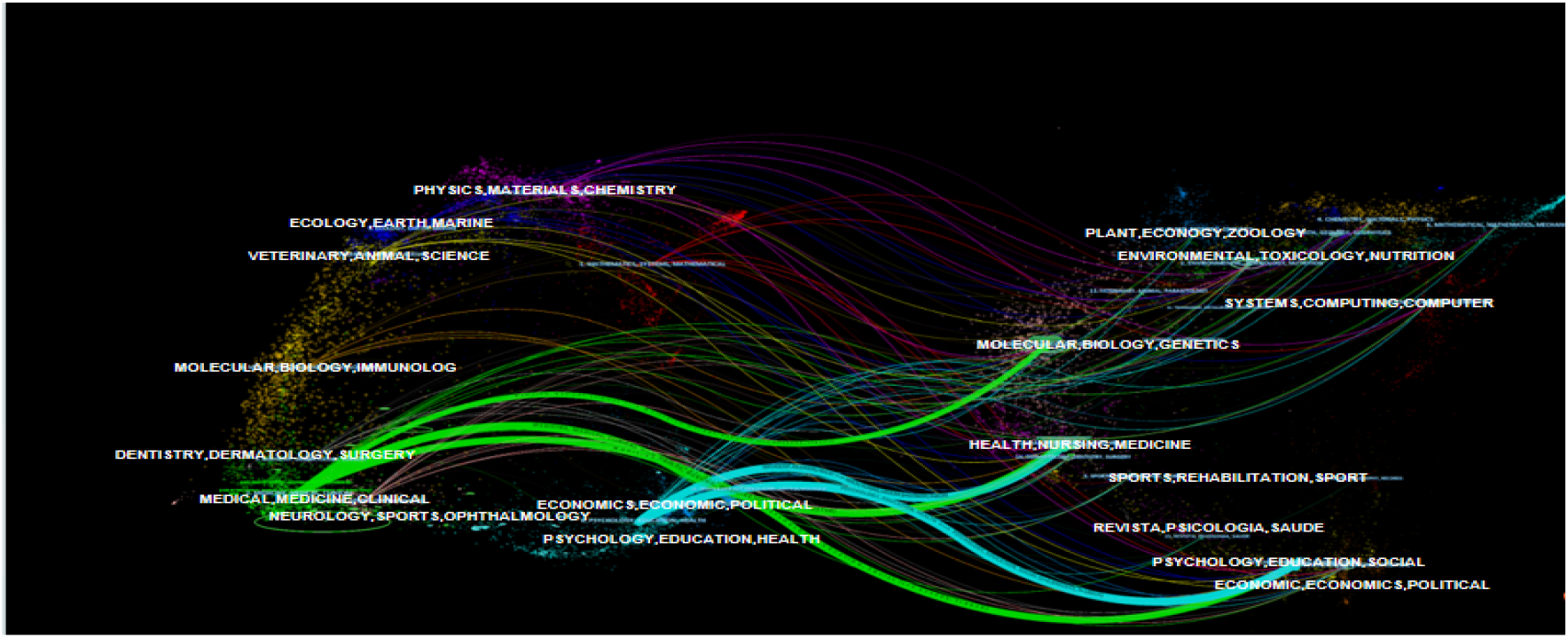
Overlay map of journals.

### 3.6 Analysis of keywords

#### 3.6.1 Analysis of keyword co-occurrence

High frequency keywords are highly condensed topics in the literature, often considered as the field’s focus. The intermediate centrality of these keywords can measure their transitional role in the literature. Table 4 lists the top ten terms with the most occurrences. The most prevalent occurrence was care (frequency: 709), followed by disaster, impact, and mental health. Fig 6 is an analysis graph of the keyword co-occurrence cooperation network. The size of nodes shows the frequency of keyword occurrence. The larger the node, the more keyword occurrences there are. According to the setting of the research method, the result is 338 nodes and 1584 lines, and the network density is 0.0278.

**Table 4.** Top 10 reference keywords related to disaster nursing.

| Rank | Keywords | Count | Centrality |
| --- | --- | --- | --- |
| 1 | care | 709 | 0.01 |
| 2 | disaster | 619 | 0.01 |
| 3 | impact | 423 | 0.01 |
| 4 | mental health | 415 | 0.02 |
| 5 | health | 371 | 0.01 |
| 6 | preparedness | 326 | 0.01 |
| 7 | natural disaster | 321 | 0.01 |
| 8 | disaster preparedness | 324 | 0.00 |
| 9 | management | 290 | 0.02 |
| 10 | post traumatic stress disorder | 265 | 0.02 |

**Fig 6.**
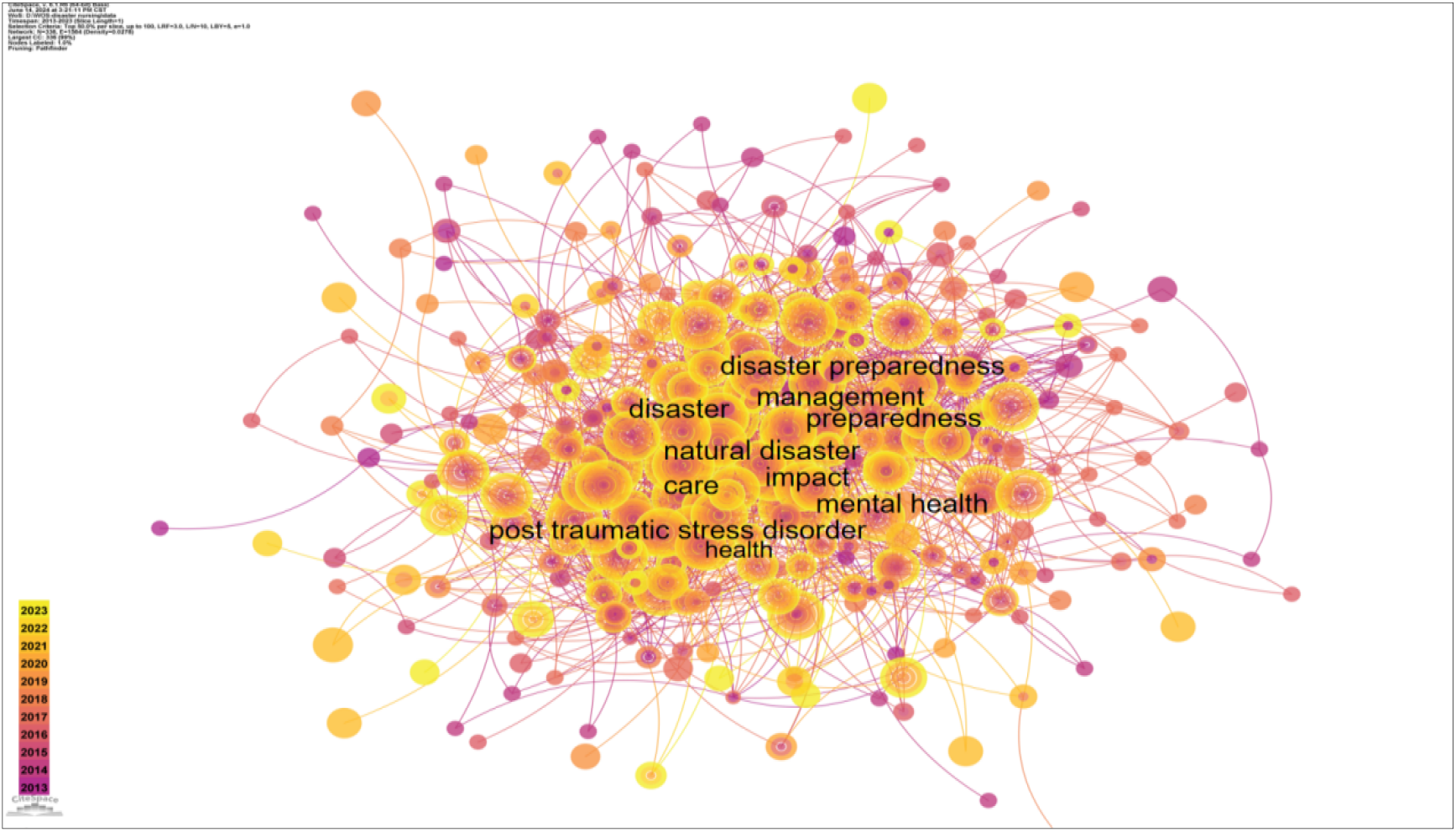
Map of keywords.

#### 3.6.2 Analysis of keyword clusters

Cluster labels reflect the overall research structure of disaster nursing. This study used keyword cluster analysis to understand the current research hotspots in disaster nursing. If the average contour value S is greater than 0.5, the clustering result is considered reasonable. The clustering module value Q is crucial for evaluating rationality indicators. When Q is greater than 0.3, the cluster community structure is significant [2]. According to the results of Q (Q = 0.4051) and S (S = 0.6926) of the mapped keyword clusters, **Fig 7** demonstrates that the clusters are convincing and helpful in identifying research hotspots in the field of disaster nursing. Summarizing the characteristics of these clusters allows for their division into three distinct topics.

**Fig 7.**
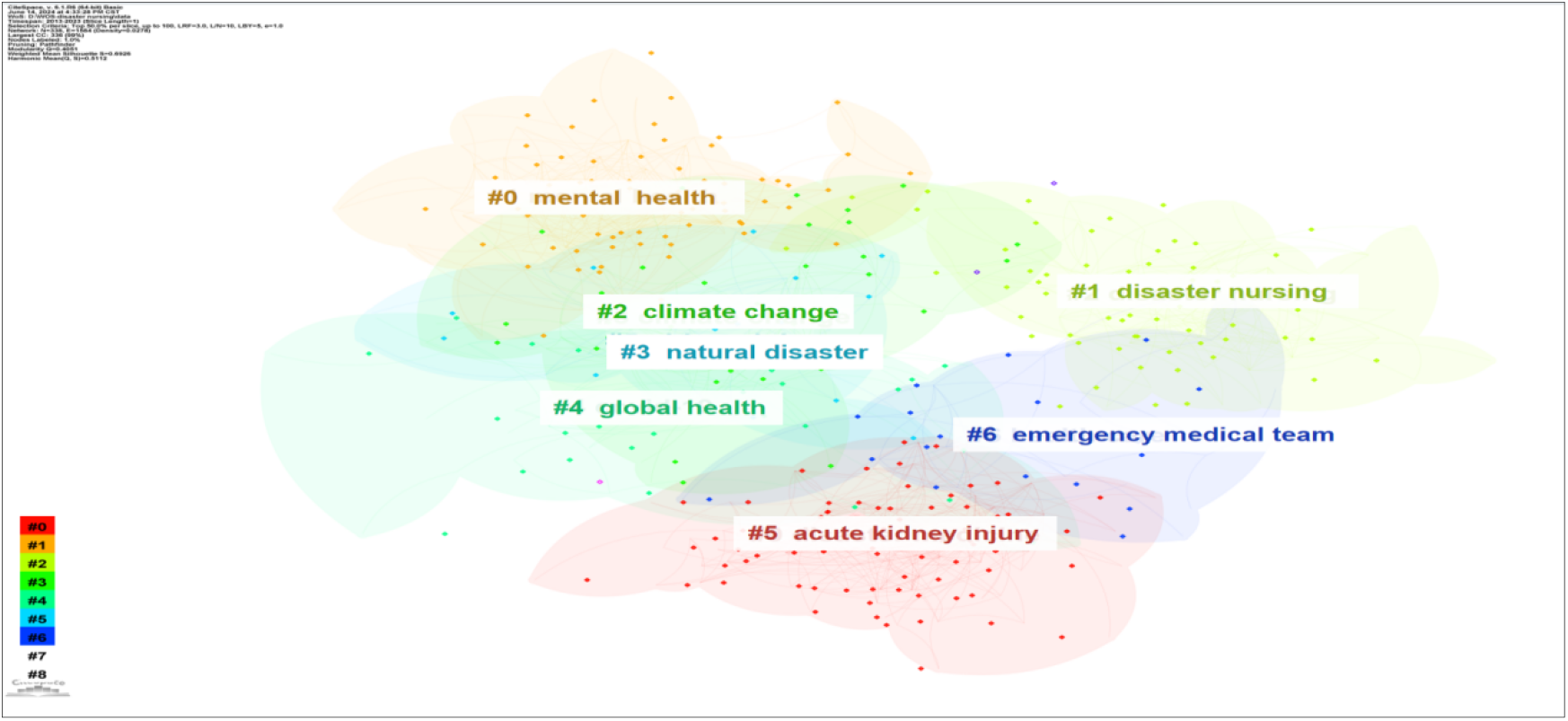
Map of keyword clusters.

##### 3.6.2.1 disaster rescue

The research hotspots in disaster rescue primarily concentrate on the evaluation, classification, treatment, and first aid of injured individuals. Disaster rescue is a crucial part of disaster nursing, which involves the rapid response and effective treatment of the affected people. The primary task in the disaster rescue process is to assess and classify the injured, which helps to allocate medical resources reasonably and improve treatment efficiency. With the continuous development and improvement of medical technology, many scholars and experts are exploring more advanced methods for the assessment and classification of the wounded, so as to more accurately judge the condition and treatment needs of the wounded. Through the analysis of the wound type, location, deep tissue bacteria culture, and other data of earthquake victims rescued from collapsed buildings after the earthquake, it was found that the most common wound type was abrasions, followed by necrotic wounds caused by crushing. Wound and skin disinfection, debridement, and negative pressure wound treatment (NPWT) are the most commonly used wound care methods [3].To reduce the risk of human casualties, a new robust mathematical model can simultaneously locate and allocate healthcare facilities, including different levels of medical care services, taking into account the characteristics of normal and disaster situations. Furthermore, the model can facilitate the emergency provision of basic services for temporary outpatient centers.[4].

In addition, the upgrading of first aid technology and equipment also provides more options and possibilities for disaster rescue. For example, FTB (Fast Triage in Burns) is a simple, quick, and credible means of segregating burn victims. It is based on parameters that can be easily evaluated without additional equipment and is dedicated to use in pre-hospital care during mass casualty events, both in civilian and battlefield circumstances [5]. Furthermore, it is imperative that disaster relief efforts stress the collaboration and effective exchange of information among team members. Rescue professionals at the disaster site must collaborate closely and acquire prompt catastrophe information and resource assistance to ensure that the injured individuals receive fast and efficient treatment and can better manage the rescue efforts.

##### 3.6.2.2 disaster management

Research on disaster management focuses on disaster early warning and preparedness, post-disaster assessment, and rehabilitation and reconstruction. Effective disaster management helps reduce disaster losses and protect people’s lives and property. In the pre-disaster early warning stage, disaster management departments need to pay close attention to meteorological, geological, and other disaster early warning information, timely issue early warning notices, and remind the public to take preventive measures. Furthermore, disaster management departments must formulate emergency plans to clarify each department’s responsibilities and tasks so that they can respond quickly when disasters occur. Many papers have been published on the cross-sectional survey and related training of nurses’ disaster preparedness. These include the cross-sectional survey of nurses’ disaster preparedness in operating rooms [6], emergency department[7], and general nurses [8]. These studies aim to understand the level of nurses’ disaster knowledge, skills, experience, and other aspects. However, there is a dearth of literature on how to improve nurses’ disaster preparedness interventions, and future research on this topic needs to be strengthened. Post-disaster assessment is an important link in disaster management; through disaster loss assessment, scope, etc., it can provide a scientific basis for post-earthquake recovery and reconstruction. In the recovery and reconstruction phases, disaster management departments need to coordinate resources and organize forces for post-disaster reconstruction as soon as possible to restore the normal production and living order of the affected areas. Information technology and big data are becoming more widely used in disaster management processes. Satellite remote sensing, unmanned aerial vehicles, and other technical means enable real-time monitoring and data analysis at the disaster site, thereby providing more accurate and timely information support for disaster management. At the same time, the mining and analysis of big data can also help disaster management departments better predict disaster trends and formulate more effective prevention and response measures. Saudi Arabia’s health sector has used drones to reduce disaster response times, increase access to underserved areas, and reduce the burden on existing medical infrastructure [9]. The successful application of drones in disaster response and pre-hospital care, which improves patient treatment outcomes, efficiency, and cost savings, has become an innovative method for healthcare services.

##### 3.6.2.3 disaster impact

Natural disasters are unpredictable, and large-scale disasters such as COVID-19, earthquakes, hurricanes, and tsunamis have long-term social, economic, and environmental impacts on affected areas that have caused great harm to human beings. Terrorism, as a highly destructive act, can also have a profound impact on society, the economy, and international relations. The number of terrorist attacks around the world is increasing, and the shooting and explosive circumstance complex, are highly dangerous to rescue forces and subsequent hospital medical. The European Union, in its efforts to combat terrorism, launched the Horror and Disaster Surgical Nursing (TDSC (R)) courses, which aim to equip clinical decision-makers with the necessary skills to tackle terror and disaster challenges.

Disaster also impacts the physical and psychological health of refugees, disaster survivors, and emergency care workers, including their daily behavior, physiological indicators, and mental states, such as distress syndrome and post-traumatic stress disorder(PTSD). Previous studies mostly focused on the impact of disasters on the physical and mental health of affected people, such as psychological trauma and disease transmission. Research has now drawn attention to the psychological issues faced by medical staff during the earthquake response. Their exposure to horrific scenes, including the sight of dead bodies and severely injured people, exposed them to numerous mental health consequences. They have a higher short- and long-term risk of developing PTSD. Therefore, the mental health of medical workers involved in disaster relief needs attention, and it is best to receive training on stress management, psychological resilience, and how to express their feelings and emotions before and after a disaster [10].It is interesting to note that some scholars foundacupuncture is a non-psychological and non-pharmacological intervention, as an alternative,complementary and integrative medicine, it may help overcome barriers to PTSD [11]. Besides, post-disaster reconstruction can provide guidance in health education and disease prevention, so as to promote the sustainable development of the affected areas, which has always been a research hotspot in disaster nursing. In general, the reconstruction and restoration of emergency services can take years. Because reconstruction resources are limited, decision-makers cannot reconstruct all hospitals at the same time. Decision-makers frequently have to prioritize the reconstruction sequence, resulting in a poorly planned process. There are scholars who have modeled emergency services as an M/M/s queuing system that takes into account the priority treatment of critically ill patients and has developed a greedy algorithm to plan an efficient reconstruction of the healthcare system. This system can shorten treatment time compared to commonly implemented policies and more than 3-fold reduce the number of untreated patients during reconstruction in a worst-case scenario (70% of hospital capacity disrupted), informing decision-makers about future post-earthquake reconstruction plans [12].

#### 3.6.3 Analysis of variation in clusters

Fig 8 display the results of timeline and landscape view analyses, which demonstrate the evolution of popular research topics over time and aid in understanding the field’s evolution. For example, the largest cluster in our study, cluster #0, indicates that mental health is the main focus of academic circles. Under this theme, research has focused on changes from PTSD to trauma, long-term real-time stress responses in children, and young people’mental health. [13]. Cluster #1 shows that disaster nursing has shifted from the survey of disaster preparedness and core competence to the research of disaster knowledge training, skill training, and pre-hospital nursing in the past 10 years, and disaster preparedness has shifted to home-based emergency preparedness for families to reduce losses [14].

**Fig 8.**
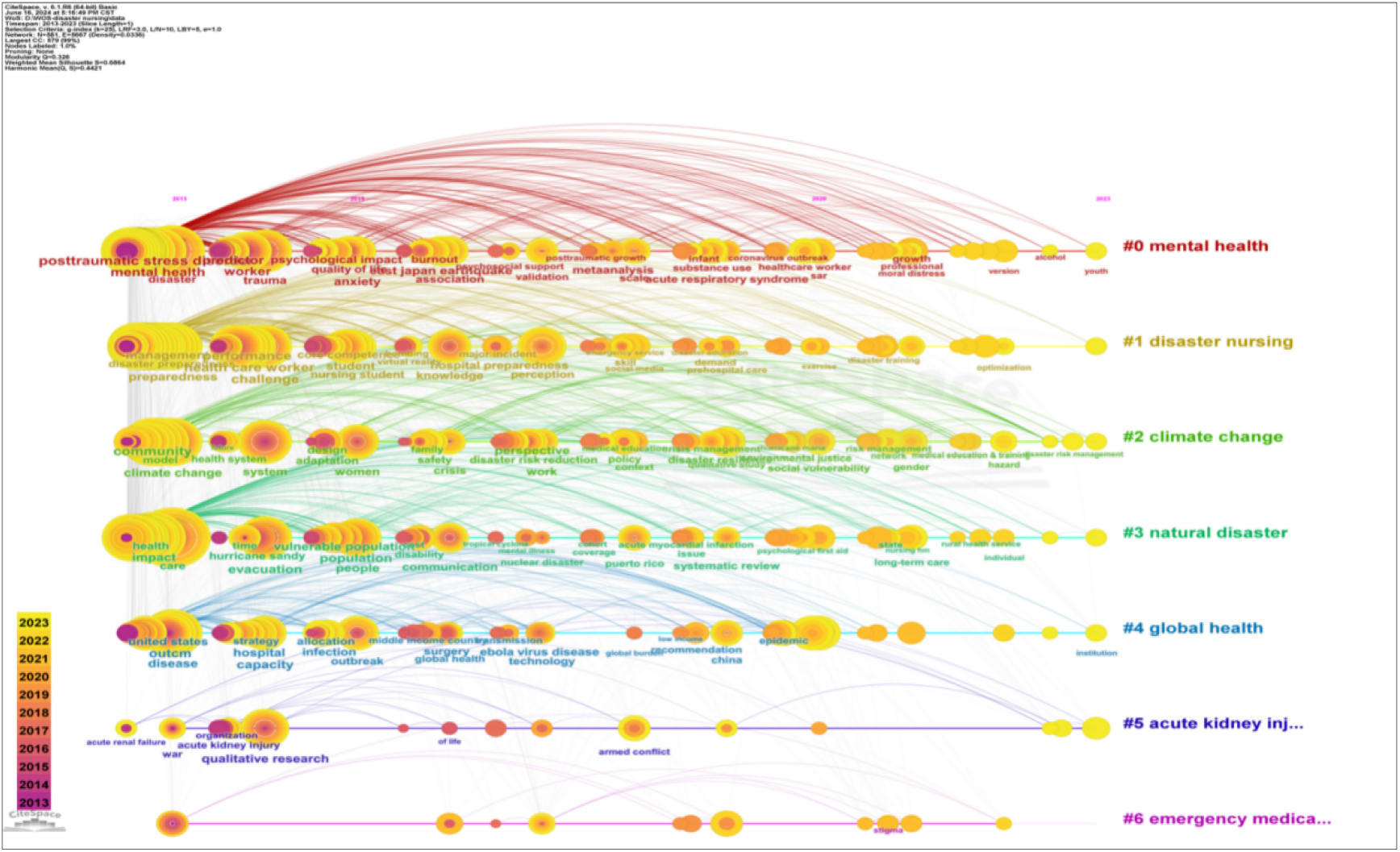
Timeline view of the keywords co-ocurrence map.

#### 3.6.4 Keywords with citation bursts

The keyword bursts can respond to trends and frontier research in the field of research. The blue line represents the time interval, while the red line represents the interval period with the keyword burst [15]. From 2013 to 2018, Hurricane Katrina has been growing rapidly. This indicates that the Hurricane Katrina event generated a lot of research attention during this time. It has not only caused huge loss of life and property, but also exposed the challenges and inadequacies of disaster care in responding to extreme weather events. In addition, other keywords such as “disease outbreak,” “nuclear disaster,” and “emergency service” also showed a clear upward trend. The growth of these keywords in the field of disaster nursing research is also expanding and deepening, from the simple disaster relief and nursing to a wider range of fields, such as post-disaster psychological rehabilitation, optimization of disaster response mechanisms, and promotion of disaster relief efficiency. It can be seen that with the intensification of global climate change and the frequent occurrence of extreme weather events, the field of disaster care is facing unprecedented challenges. How to better solve these problems and improve the level of disaster care will be the focus of future research.

**Fig 9.**
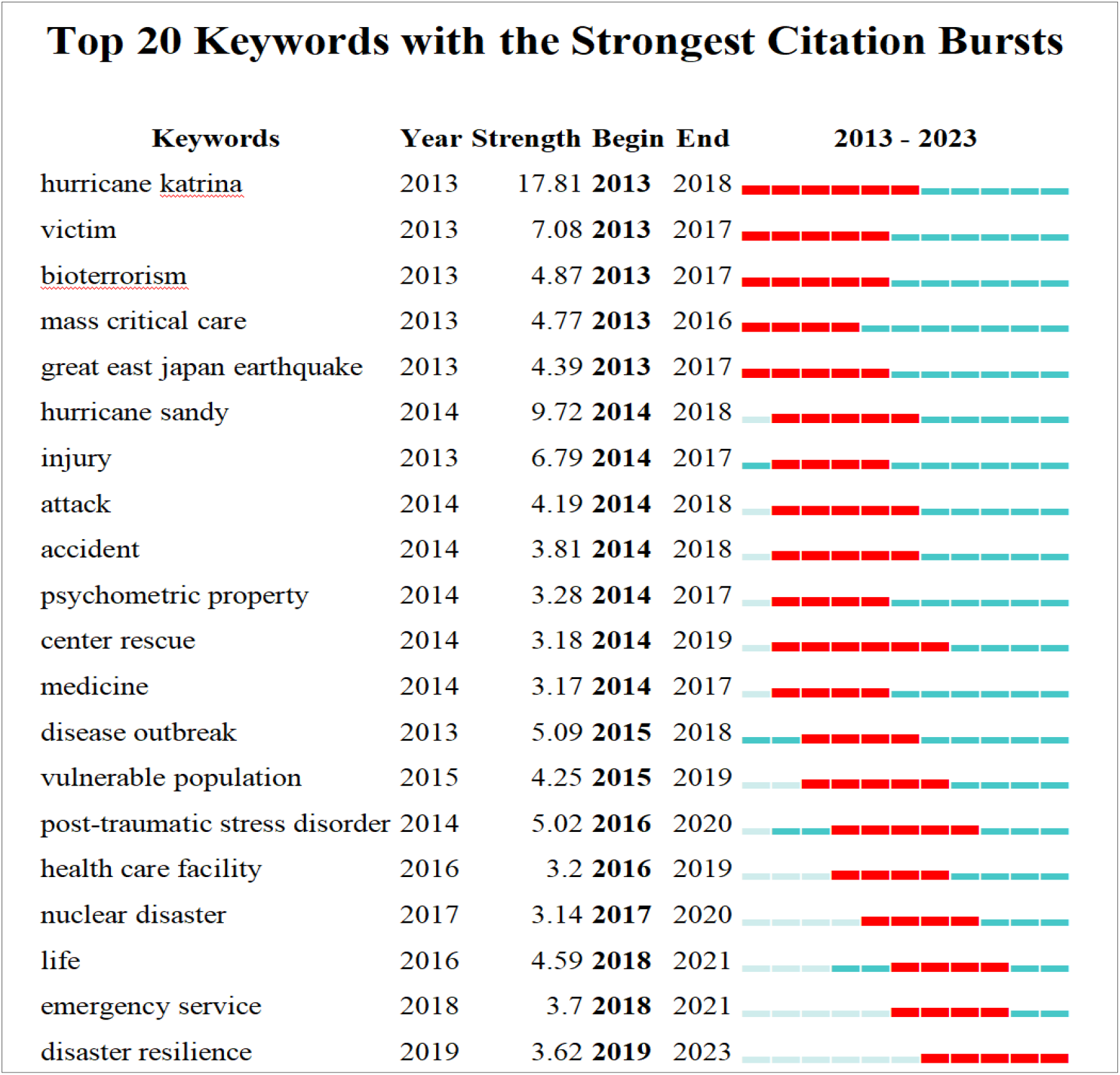
Top 20 keywords with the strongest citation bursts.

## 4. Discussion

### 4.1 Evolution and deepening of disaster nursing research hotspots

Nearly a decade of disaster nursing has identified an overall rising trend, indicating that the research field deserves researchers’ attention. According to Fig 1’s national distribution, research on disaster nursing is primarily concentrated in developed countries. The United States, Australia, Japan, the United Kingdom, and China have the top five publications, with the United States having the largest number. The United States holds six positions among the top 10 institutions, demonstrating its leadership in this field of research as well as its significant role in its development. The CiteSpace visualization analysis reveals that the hotspots of disaster nursing research has evolved and deepened over time. It has expanded from its initial focus on disaster rescue and nursing to include disaster response mechanism optimization, disaster preparedness, disaster resilience and disaster nursing education.

#### 4.1.1 Disaster response and preparedness

Studies reveal that over the past 20 years, 430 terrorist attacks have targeted hospitals worldwide, resulting in 1,291 deaths and 1,921 injuries [16]. These disasters make more and more scholars pay close attention to how to handle extreme events such as terror attacks, guarantee the safe operation of medical facilities, and improve disaster response capacity. In recent years, with the frequent occurrence of disaster events, research on disaster nursing has increasingly focused on enhancing the efficiency and quality of coping with challenges. After Hurricane Katrina swept across the United States, the fragility of the medical system was fully exposed [17]. This reveals the deficiency of disaster nursing in response to extreme weather disasters but also promotes research in the field of disasters, but promotes research in the field of disaster nursing to deeper development and disaster preparation, which has become the focus of researchers. At this stage of disaster nursing research, which started with the government, hospitals, and medical personnel, medical personnel gradually turned to families as the unit of vulnerable groups emergency preparedness education [18].

#### 4.1.2 Disaster resilience

The huge losses caused by disasters make the concept of disaster resilience has gained increasing attention in disaster nursing research. Disaster resilience refers to the ability of individuals, communities, and systems to recover from disasters and adapt to changing conditions. In disaster nursing, resilience is seen as a key factor in promoting long-term recovery and reducing the impact of disasters on health and well-being.Recent research has focused on strategies to enhance disaster resilience at various levels. At the individual level, this includes promoting self-care, stress management, and coping skills. At the community level, resilience-building efforts aim to strengthen social networks, promote community participation, and develop disaster preparedness plans. At the system level, disaster resilience are supported through government policies to reduce disaster losses

Disaster resilience often focuses on repairing facilities, but psychological issues are often overlooked. The trauma, anxiety, and depression that survivors experience can be long-lasting, affecting not only their individual well-being but also their ability to function in society. Recent disaster nursing research begins to pay attention to this. It has emphasized the need for comprehensive mental health support systems. The focus on the mental health issues faced by disaster nurses not only broadens the scope of disaster nursing research, but also reflects the ongoing refinement and deepening of the disaster resilience. The study’s results indicate a correlation between disaster resilience and cluster #0 mental health, as well as the frequently occurring keywords “mental health” and “post-traumatic stress disorder.” Since nurses have the closest contact with patients among professionals, they should not only possess emergency rescue skills but also be capable of post-disaster reconstruction. For example, nurses provide psychosocial support for the negative emotions of disaster victims, such as anxiety, depression [19]. Simultaneously, rescue workers experience significant psychological strain due to the overwhelming number of deceased and injured individuals, these individuals’ psychological problem should be attentioned [20]. The COVID-19 pandemic has further highlighted the importance of mental health support in disaster nursing. Fear, and uncertainty surrounding the virus have led to a surge in mental health issues, particularly among frontline healthcare workers and those most affected by the pandemic. This has prompted researchers to explore innovative ways to provide mental health support remotely, using technologies such as telehealth and virtual reality. The integration of mental health support into disaster nursing plans is crucial for promoting disaster resilience. It is not only about treating immediate psychological trauma but also about preventing future mental health issues and building resilience among survivors.

#### 4.1.3 Disaster nursing education

In order to improve disaster response capacity and ensure people’s safety, countries have strengthened disaster nurses’ education and training. A group of professionals with disaster nursing knowledge and skills were trained by setting up relevant courses and holding training classes. To further advance this field, we can consider incorporating artificial intelligence (AI) into disaster nursing simulations. AI-driven simulations can offer nurses an unparalleled level of realism, allowing them to encounter a wider range of unpredictable scenarios. The AI system can adapt based on the nurses’ responses, providing real-time feedback and suggestions for improvement.Moreover, the integration of virtual reality (VR) technology can take the learning experience to a whole new level. Through VR, nurses can be fully immersed in disaster scenarios, feeling as if they are truly there. This sense of presence not only enhances their learning, but also prepares them psychologically for the rigors of real-life disasters. This approach not only enhances their emergency response and comprehensive skills during disasters, but also fosters improved communication and cooperation among various departments [21]. The combined application of technology and education is helpful to improve the efficiency and quality of disaster nursing education. It presents a fresh opportunity for the advancement of the disaster nursing field. It also opens up additional avenues for future research.

### 4.2 The combination of disaster nursing and intelligent technology is a research trend in the future

With the rapid development of information technology, disaster nursing research is gradually moving in the direction of digitalization and intelligence. The researchers used data and technical means such as artificial intelligence, data mining, and analysis of disasters to reveal the rules and character of disaster nursing and provide a scientific basis for disaster relief. For example, a portable ultrasound diagnosis system equipped with an artificial intelligence robotic arm can maximize the pre-screening and classification of batch casualties and provide a reliable basis for the classification and evacuation of batch casualties at the disaster accident site. Additionally, portable ultrasound can guide procedures such as endotracheal intubation, pericardial cavity puncture, and chest puncture during emergency operations, thereby improving surgical technique accuracy and expediting the process [22]. The application of these techniques helps to improve the efficiency and quality of disaster nursing.

### 4.3 Obstructions and prospects

Disaster nursing is an important part of disaster emergency management, which deserves close attention from the whole society. Although the research in the field of disaster nursing has made some progress, the development of disaster nursing in various countries is not balanced, and it still faces many challenges in practice. On the one hand, the suddenness and uncertainty of disaster events have brought significant challenges to disaster nursing, which requires nursing staff to have a high degree of adaptability and professional quality. On the other hand, the limitations and uneven distribution of disaster nursing resources have also restricted the effective implementation of disaster nursing. Future disaster nursing research, therefore, needs to pay more attention to practical application, strengthen international cooperation, promote interdisciplinary collaboration and innovation, and give full play to the role of policy guidance and support to promote the prosperity and development of disaster nursing careers.

## 5. Limitations

This research to retrieve only the Web of Science database literature in the future can expand the retrieval scope, fully displaying the nurse disaster readiness study track and trend.

## 6. Conclusions

This study used CiteSpace software to analyze the literature related to disaster nursing over the past 10 years and found that the United States was in a leading position in this field. Training and education in disaster nursing, disaster resilience, and emergency preparedness for natural disasters, bioterrorism, and public health events were research hotspots. Artificial intelligence, big data, integration, and other new concepts will become the trend of disaster nursing research in the future. Nurses are the largest group in the medical system, as well as the largest group in disaster rescue. They should constantly learn disaster nursing knowledge and skills to better cope with the growing risk of disasters.

## Data Availability

The data comes from the Web of Science Core Collection.

https://clarivate.com.cn/solutions/web-of-science/

## Acknowledgments

We appreciate the data availability via the Web of Science Database as well as the cooperation of the authors.

## Author contributions

Conceptualization: Haixia Xie, Zhi Zhang.

Data curation: Haixia Xie, Zhi Zhang, Fulan Li, Tianshuang Yu.

Formal analysis: Haixia Xie.

Funding acquisition: Ruijuan Han.

Resources: Zhi Zhang, Fulan Li.

Supervision: Ruijuan Han.

Visulazation: Haixia Xie.

Writing – original draft: Haixia Xie.

Writing – review & editing: Haixia Xie, Zhi Zhang, Ruijuan Han.

## References

1. Fu Y, Zhao J, Zhang W, Du H, Cao Z, Chen X. Global research trends in sexual health care: A bibliometric and visualized study. J Clin Nurs. 2024;33(2):422–31. 10.1111/jocn.16915

2. Chen C. CiteSpace II: detecting and visualizing emerging trends and transient patterns in scientific literature. J Am Soc Inf Sci Technol. 2006;57:359–77. 10.1002/asi.20317

3. Ulusoy S, Kilinç I, Oruç M, Özdemir B, Ergani HM, Keskin Ö, et al. Analysis of wound types and wound care methods after the 2023 Kahramanmaras earthquake. JOINT DISEAS ES AND RELATED SURGERY. 2023;34(2):488–96. 10.52312/jdrs.2023.1128

4. Alinaghian M, Hejazi SR, Bajoul N, Velni KS. A novel robust model for location-allocation of healthcare facilities considering pre-disaster and post-disaster characteristics. SCIENTIA IRANICA. 2023;30(2):619–41. 10.24200/sci.2021.5743.1459

5. Surowiecka-Pastewka A, Witkowski W, Kawecki M. A New Triage Method for Burn Disasters:Fast Triage in Burns (FTB). MEDICAL SCIENCE MONITOR. 2018;24:1894–901. 10.12659/MSM.905197

6. Rostami M, Babajani-Vafsi S, Ziapour A, Abbasian K, Mohammadimehr M, Zareiyan A. Experiences of operating room nurses in disaster preparedness of a great disaster in Iran: a qualitative study. BMC emergency medicine. 2023;23(1). 10.1186/s12873-023-00903-w

7. Zhang JE, Yang L, Cao X, Ren Y, Han X, Zang ST, et al. Assessment of disaster preparedness and related impact factors among emergency nurses in tertiary hospitals: descriptive cross-sectional study from Henan Province of China. Frontiers in public health. 2023;11. 10.3389/fpubh.2023.1093959

8. Ediz Ç, Yanik D. Disaster preparedness perception, pyschological resiliences and empathy levels of nurses after 2023 Great Turkiye earthquake: Are nurses prepared for disasters: A risk management study. PUBLIC HEALTH NURSING. 2024;41(1):164–74. 10.1111/phn.13267

9. Al-Wathinani AM, Alhallaf MA, Borowska-Stefanska M, Wisniewski S, Sultan MAS, Sam man OY, et al. Elevating Healthcare: Rapid Literature Review on Drone Applications for Streamlining Disaster Management and Prehospital Care in Saudi Arabia. HEALTHCARE. 2023;11(11). 10.3390/healthcare11111575

10. Tahernejad S, Ghaffari S, Ariza-Montes A, Wesemann U, Farahmandnia H, Sahebi A. Post-traumatic stress disorder in medical workers involved in earthquake response: A systematic review and meta-analysis. Heliyon. 2023;9(1). 10.1016/j.heliyon.2023.e12794

11. Seung HB, Leem J, Kwak HY, Kwon CY, Kim SH. Acupuncture for military veterans wi th posttraumatic stress disorder and related symptoms after combat exposure: Protocol for a scoping review of clinical studies. PLOS ONE. 2023;18(4). 10.1371/journal.pone.0273131

12. Alisjahbana I, Ceferino L, Kiremidjian A. Prioritized reconstruction of healthcare facilities after earthquakes based on recovery of emergency services. RISK ANALYSIS. 2023;43(9): 1763–78. 10.1111/risa.14076

13. Witt A, Sachser C, Fegert JM. Scoping review on trauma and recovery in youth after nat ural disasters: what Europe can learn from natural disasters around the world. EUROPEAN CHILD & ADOLESCENT PSYCHIATRY. 2024;33(3):651–65. 10.1007/s00787-022-01983-y

14. Griffin JS, Hipper TJ, Chernak E, Kurapati P, Lege-Matsuura J, Popek L, et al. Home-Based Emergency Preparedness for Families of Children and Youth With Special Healthcare Needs: A Scoping Review. Health security. 2023;21(3):193–206. 10.1089/hs.2022.0119

15. Chen X, Chen X, Lai Y. Development and emerging trends of drug resistance mutations i n HIV: a bibliometric analysis based on CiteSpace. Frontiers in microbiology. 2024;15:137 4582. 10.3389/fmicb.2024.1374582

16. McNeilly B, Jasani G, Cavaliere G, Alfalasi R, Lawner B. The Rising Threat of Terrorist Attacks Against Hospitals. Prehospital and disaster medicine. 2022;37(2):223–9. 10.1017/S1049023X22000413

17. Hanfling D, Bouri N. Foreign Medical Teams: What Role Can They Play in Response to a Catastrophic Disaster in the US? Disaster medicine and public health preparedness. 2013 ;7(6):555–62. 10.1017/dmp.2013.95

18. Shang JJ, Chastain AM, Perera UGE, Quigley DD, Fu CJ, Dick AW, et al. COVID-19 Preparedness in US Home Health Care Agencies. Journal Of The American Medic Al Directors Association. 2020;21(7):924–7. 10.1016/j.jamda.2020.06.002

19. Stukova M, Cardona G, Tormos A, Vega A, Burgos G, Inostroza-Nieves Y, et al. Mental health and associated risk factors of Puerto Rico Post-Hurricane Maria. Social psychiatry a nd psychiatric epidemiology. 2023;58(7):1055–63. 10.1007/s00127-023-02458-4

20. Uddin H, Hasan MK, Castro-Delgado R. Effects of mass casualty incidents on anxiety, depression and PTSD among doctors and nurses: a systematic review protocol. BMJ OPEN. 2023;13(9). 10.1136/bmjopen-2023-075478

21. Ghahremani M, Rooddehghan Z, Varaei S, Haghani S. Knowledge and practice of nursing students regarding bioterrorism and emergency preparedness: comparison of the effects of simulations and workshop. BMC nursing. 2022;21(1). 10.1186/s12912-022-00917-y

22. Gao X, Lv Q, Hou SK. Progress in the Application of Portable Ultrasound Combined wit h Artificial Intelligence in Pre-Hospital Emergency and Disaster Sites. DIAGNOSTICS. 2023;13(21). 10.3390/diagnostics13213388

